# Student self-assessment: feasibility, advantages and limitations Example of a workshop for trainee surgeons using a suture score

**DOI:** 10.1101/2024.06.04.24308019

**Authors:** Elise Riquin, Thomas Le Nerzé, Jeanne Goulin, Louis Rony, Sophie Boucher, Ludovic Martin, Françoise Schmitt

## Abstract

**Introduction:** Evaluating skills requires developing relevant grading tools that could potentially be used for self-assessments. The goal of our study was to assess the relevance of a self-assessment (SA) grid used for a skin suture exercise as compared to an evaluation carried out at the same time by an expert (EE), while also identifying any factors likely to influence it.

**Materials and methods:** Between 2018 and 2022, an assessment grid for a skin suture exercise was used during skills assessment sessions with first-year trainee surgeons. The students were given a self-assessment grid to fill out at the same time. The results of the SA and the EE in this grid were analyzed and compared, then set against the anxiety scores of the trainees based on the STAI Y-A. A comparative analysis of trainees with a tendency to over- or underestimate themselves was carried out to identify the factors that could contribute to them misjudging their skills.

**Findings:** The data for 70 trainee doctors was analyzed. The mean of the difference in score between the SA and the EE was 2 points lower for SA than EE (p < 0.001). The mean score for the STAI Y-A was 36.2 +/- 8.9 and the learner’s anxiety had no effect on the EE but did affect the SA. When learners showed low self-confidence, they underestimated themselves, but examiners also had a tendency to give them lower grades. In multivariate analysis, the less experience a learner had, the more they tended to overestimate their abilities.

**Conclusion:** Anxiety and self-confidence affect self-assessment, but external assessment does not seem to be affected by the learner’s stress levels. Formative self-reflection based on the use of SA grids should be introduced during medical studies so as to adjust the level of technical skill perceived by trainees and to make simulation-based training more effective.

## 1. Background

For more than 20 years, it has been shown that simulation-based learning has a positive impact on training students, whose skill levels when they start graduate degrees are highly unequal (Liepert *et al*., 2019). Many studies have confirmed this phenomenon within surgical specialties (Acton *et al*., 2010; Beard *et al*., 2011; Shen *et al*., 2018). That is why many faculties of medicine, including ours, now offer a seminar to new trainee surgeons before the start of the academic year to improve and harmonize their skills (Schmitt *et al*., 2022). Yet skills must be assessed using grading tools that are relevant and fair and that can be used in formative assessments during simulation-based training sessions through the evaluation of operational skills (know-how) and through summative assessments during tests such as objective structured clinical examinations (OSCEs).

In our study, we focused more specifically on the relevance to self-assessment (SA) of a grading score developed and used during seminars for trainee surgeons to test their technical skills in suturing. The score will be used as a point of comparison with the evaluation carried out at the same time by an expert (EE), while also identifying any factors that influence it.

## 2. Materials and methods

### 2.1 Description of the training undertaken

A two-day seminar before the start of the academic year has been offered to trainee surgeons at our hospital since 2013 on a voluntary basis to provide hands-on training on the basic knowledge and skills expected from a specialist junior doctor at the beginning of the program.

Between 2018 and 2022, a study was conducted to assess whether the training had any impact on improving procedural knowledge and skills in general surgery from the early days of placement (Schmitt et al., 2022). The State-Trait Anxiety Inventory (STAI Y-A) was used at the start of the seminar, MCQs assessing knowledge were used at the start and at the end of the seminar, and a formative assessment session in the form of an OSCE was carried out 3 months after the start of the placement. To assess the session workshops, grids for evaluating technical skills were created and used by the expert trainers to help during debriefing but also by the students themselves as a form of self-assessment. This study has received approval by both Angers University Hospital ethics committee and CNIL (Commission Nationale Informatique et Libertés) (N°2019-002).

### 2.2 Study objectives and endpoints

For our study, we focused on the ‘knots and sutures’ workshop performed using a synthetic skin kit (Suture Skills Trainer 3500, VATA, Medicalem, France) and covering manual surgical knots, 5 cm intradermal continuous sutures, and stop knots tied using a needle holder. The primary objective of our study was to determine how reliable self-assessments (SAs) are compared to expert evaluations (EEs) when using a multimodal assessment grid.

The secondary objectives were to examine (1) the inter-observer reliability of each sub-score in the multimodal grid to determine which one was the most reliable to include as part of a self-assessment, and (2) the factors that influence learner SA, including perceived stress using STAI Y-A and self-confidence using specific analysis of item 11 in this scale. We also examined the learner’s over- or underestimation based on the score difference between the SA and the EE.

### 2.3 Inclusion criteria and exclusion criteria

The inclusion criteria for our study were: any first-semester trainee doctor specializing in surgery or obstetrics and gynecology, taking part in the study voluntarily, enrolled in a DES (*diplôme d’études spécialisées*, post-graduate degree course) at Angers University Hospital, and having signed an informed consent form. The non-inclusion criteria were: refusal to take part in the study and any trainee doctor with previous experience in surgery (FFI or *faisant fonction d’interne* – acting as junior doctor; foreign doctor; *droit au remords* – requests for change in specialization; etc.). The exclusion criteria were: stopping the placement or changing specialization before the end of the 3rd month of the placement.

### 2.4 Assessment grids and questionnaires

Prior to this study, we had a first validation phase for the assessment grid used in the skin suture exercise, created specifically by the trainers (the data is being published at the time of writing). The multimodal assessment grid was based on a score out of 50 points and included: a task-by-task description checklist out of 20 points, a global technical performance rating scale based on the generic validated assessment OSATS (Objective Structured Assessment of Technical Skill) out of 25 points, and a speed score rating the duration of the exercise out of 5 points (Shen *et al*., 2018). The multimodal assessment grid (checklist and OSATS) can be found in Appendix 1.

To describe the differences between the self-assessments (SAs) and expert evaluations (EEs), we calculated the mean of the total difference in scores between the SA and the EE (i.e. Diff = AE score - EE score). To examine the specific characteristics of learners who under- or overestimate themselves, we defined the ‘extreme’ groups based on the point difference of the total score between the SA and the EE. We chose a deviation of ≥ 10% (i.e. 3 points or more of difference between the SA and the EE).

The STAI Y-A was used to measure the level of anxiety felt by learners and it includes 20 items designed to identify what the participant felt in the moment. Each item has a score from 1 to 4 (4 being the highest level of anxiety). The total score is the sum of the 20 questions and ranges from 20 to 80. STAI scores are classified as very high anxiety (> 65), high anxiety (56–65), moderate anxiety (46–55), low anxiety (36–45) and very low anxiety (< or = 35) (Spielberger *et al*., 1983). The scale can be used to study the impact of anxiety on cognitive performance. Item 11 (‘I feel self-confident’) was examined individually for this study (STAI Y-A self-confidence) to identify the impact of self-confidence on the learner’s self-assessment (SA). This item is reversed, and a score of 1 means high self-confidence while a score of 4 means very low self-confidence.

### 2.5 Statistical analyses

Descriptive analyses are presented for the entirety of the data collected on the participants included in the study. The qualitative variables were studied in terms of frequency and percentage in accordance with the methods of the parameter. Quantitative variables were analyzed in terms of medians, extreme values, or means and standard deviations. Only study participants with documented endpoints that can be analyzed and with a three-month full follow-up were analyzed. All the tests are bilateral and were performed with an alpha risk of 5%. Quantitative variables were compared using the Mann–Whitney U test and qualitative variables were analyzed using Fisher’s exact test. Correlation analysis was carried out using Spearman and/or Pearson correlation coefficients after checking whether the data sets were normally distributed using the Shapiro–Wilk test and expressed as a correlation coefficient (r). Factors associated with trainee doctors over- or underestimating their skills were identified through logistic regression using a stepwise model in descending order.

## 3. Findings

### Description of the cohort

Between 2018 and 2022, 101 trainee doctors specializing in surgery or gynecology were enrolled at Angers University Hospital. Among them, 18 trainee doctors were excluded from the study because they failed to attend the suture OSCE, and 13 trainee doctors were excluded because they attended the suture OSCE but did not fill in the self-assessment grid. In total, 70 trainee doctors whose data could be analyzed were included in the study. The mean age of the trainee doctors was 24.5 +/- 1.0 year. There were 36 men and 34 women (i.e. 48.6% of women). For 6 trainee doctors, their dominant hand was their left hand (8.8%). The mean cumulative duration of surgery placements during the *externat* (second, third and fourth years of the medical degree) was 34.0 +/- 20.0 weeks and the number of surgery placements during the *externat* was 4.1 +/- 1.6. The mean score for the STAI Y-A was 36.2 +/- 8.9 and the mean score for self-confidence in STAI Y-A (item 11) was 2.4 +/- 0.8. The mean of the exercise duration scores was 2.1 +/- 1.7 minutes.

### Correlation between SA and EE scores

The scores obtained for the checklist, for the OSATS and for the total score between the SA and the EE (Table 1) showed that the SA score was lower than the EE score. The mean of the total difference in scores between the SA and the EE was -2.2 +/- 4.9. For the checklist and OSATS sub-scores, the mean of the difference in points was respectively -1.1 +/- 2.6 and -1.1 +/- 3.1.

**Table 1:**
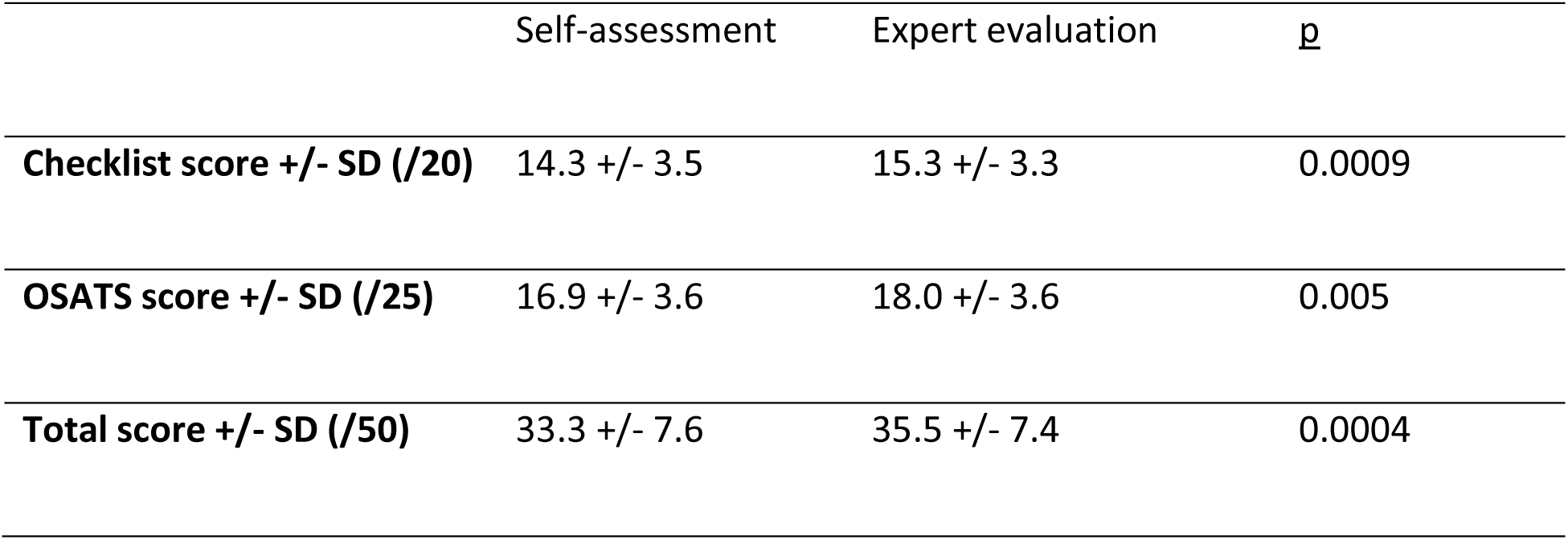
Scores by self-assessment or expert evaluation. Legend: SD: standard deviation, Total score = OSATS score + checklist score + speed score

There was a strong and significant correlation between SA and EE (Figure 1). The total score was the most correlated (r = 0.789; p < 0.0001). The checklist score was slightly more correlated between SA and EE (r = 0.715, p < 0.0001) than the OSATS score (r = 0.629, p < 0.0001).

**Figure 1:**
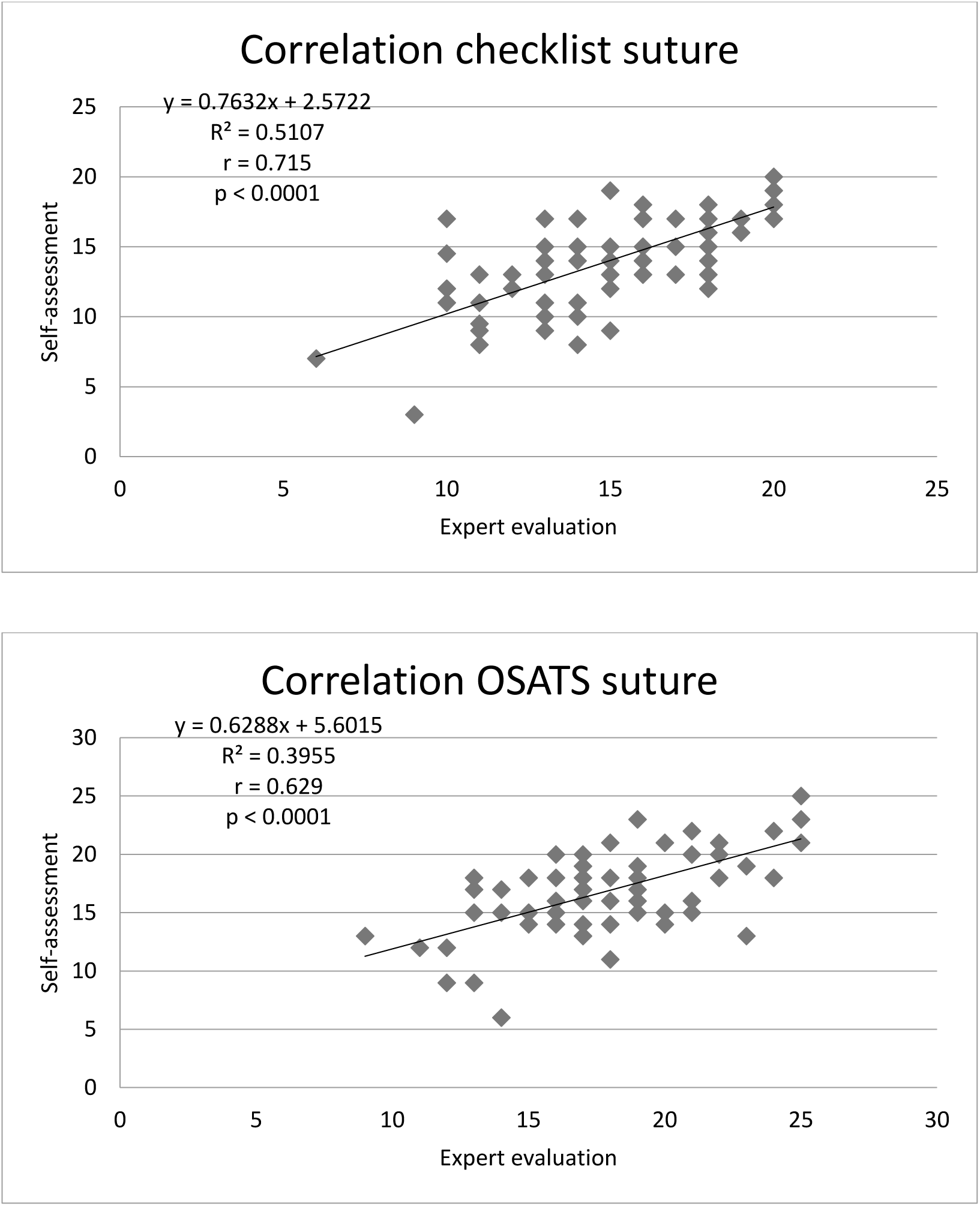
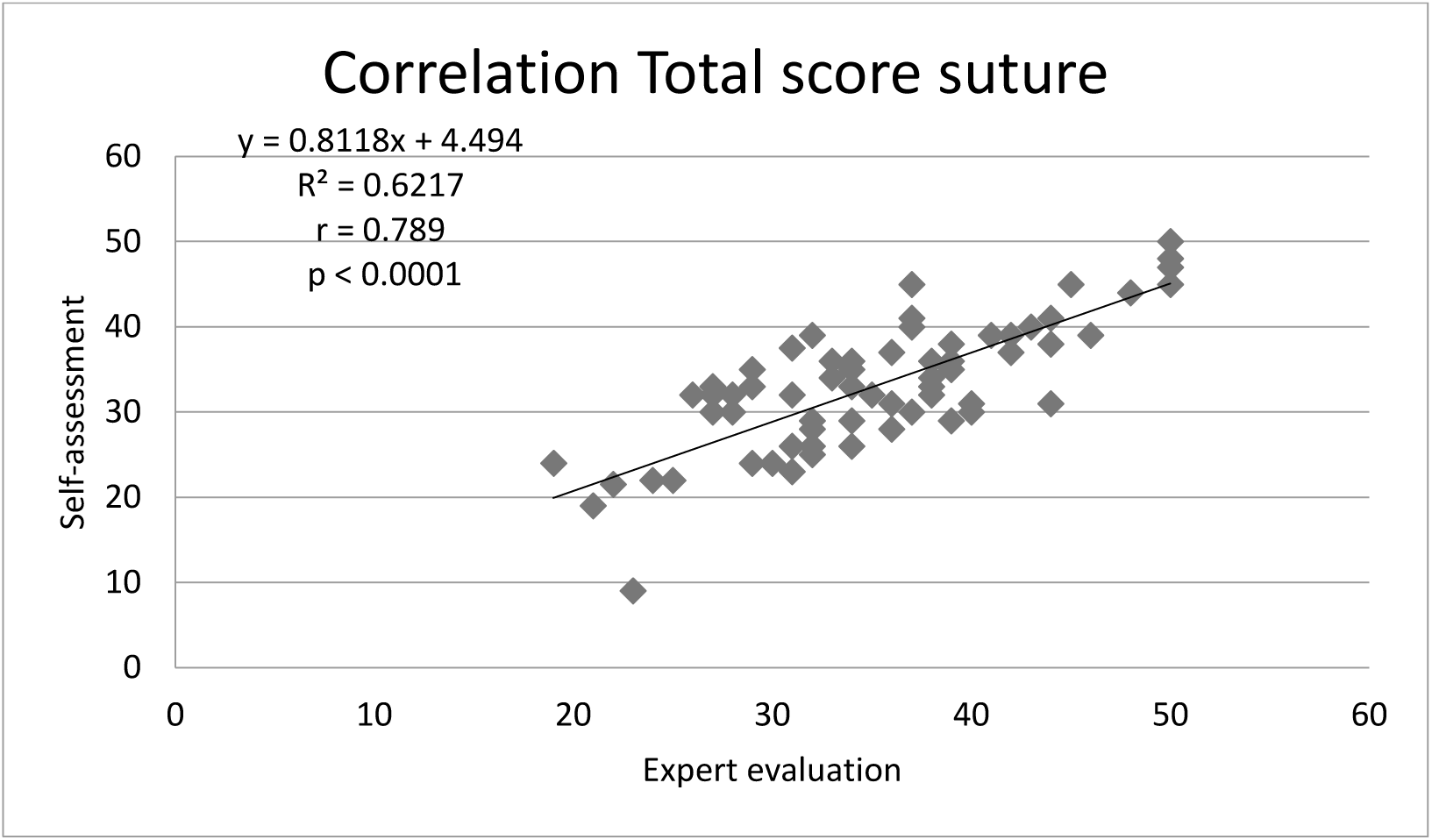
Correlation analysis EE vs SA Legend: r: correlation coefficient, R^2^: coefficient of determination

### Correlation between technical skill and stress

The results of testing the correlation between the scores of the suture grid and the anxiety score (STAI Y-A) and self-confidence score are shown in Figures 2 and 3, respectively.

**Figure 2:**
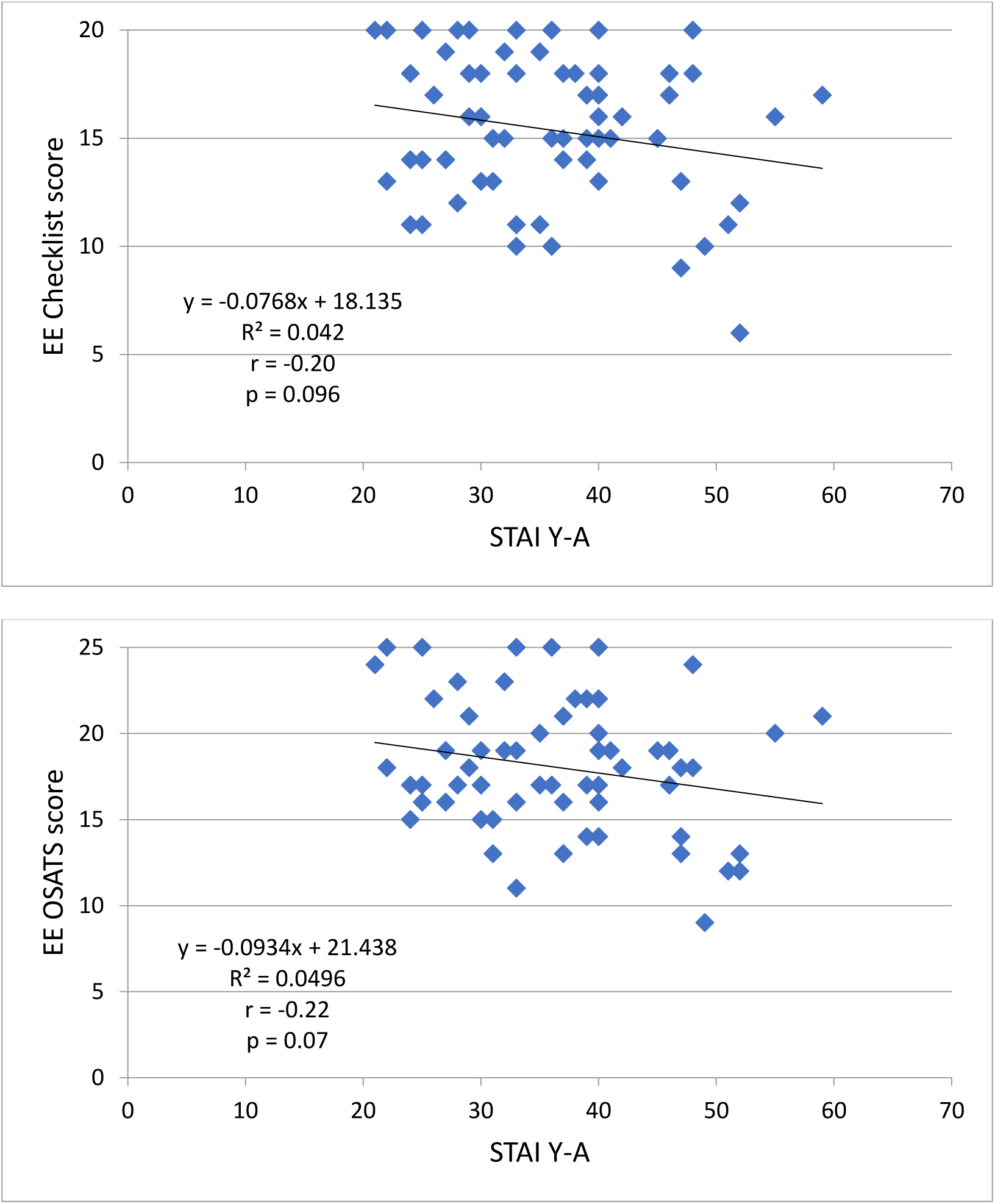
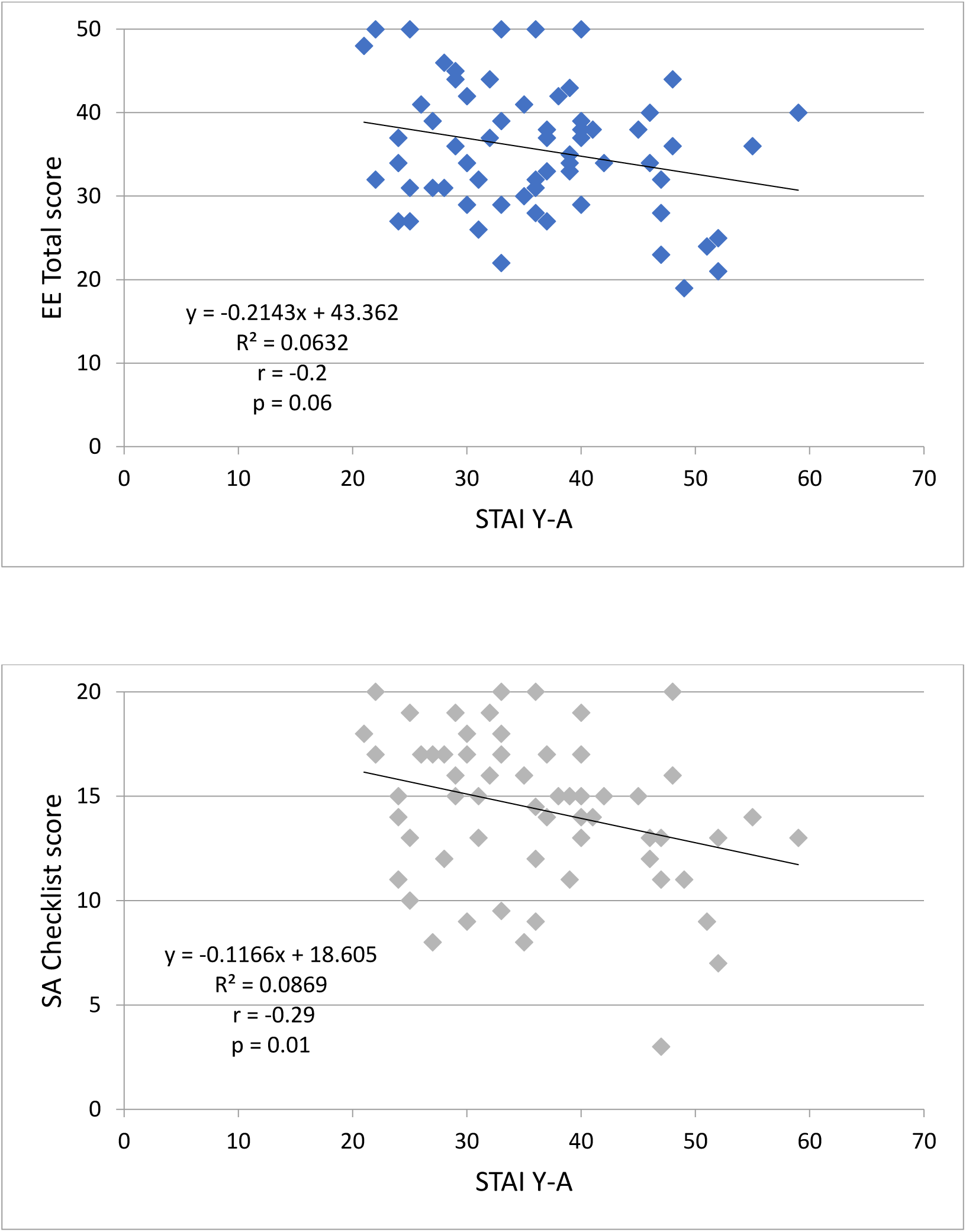
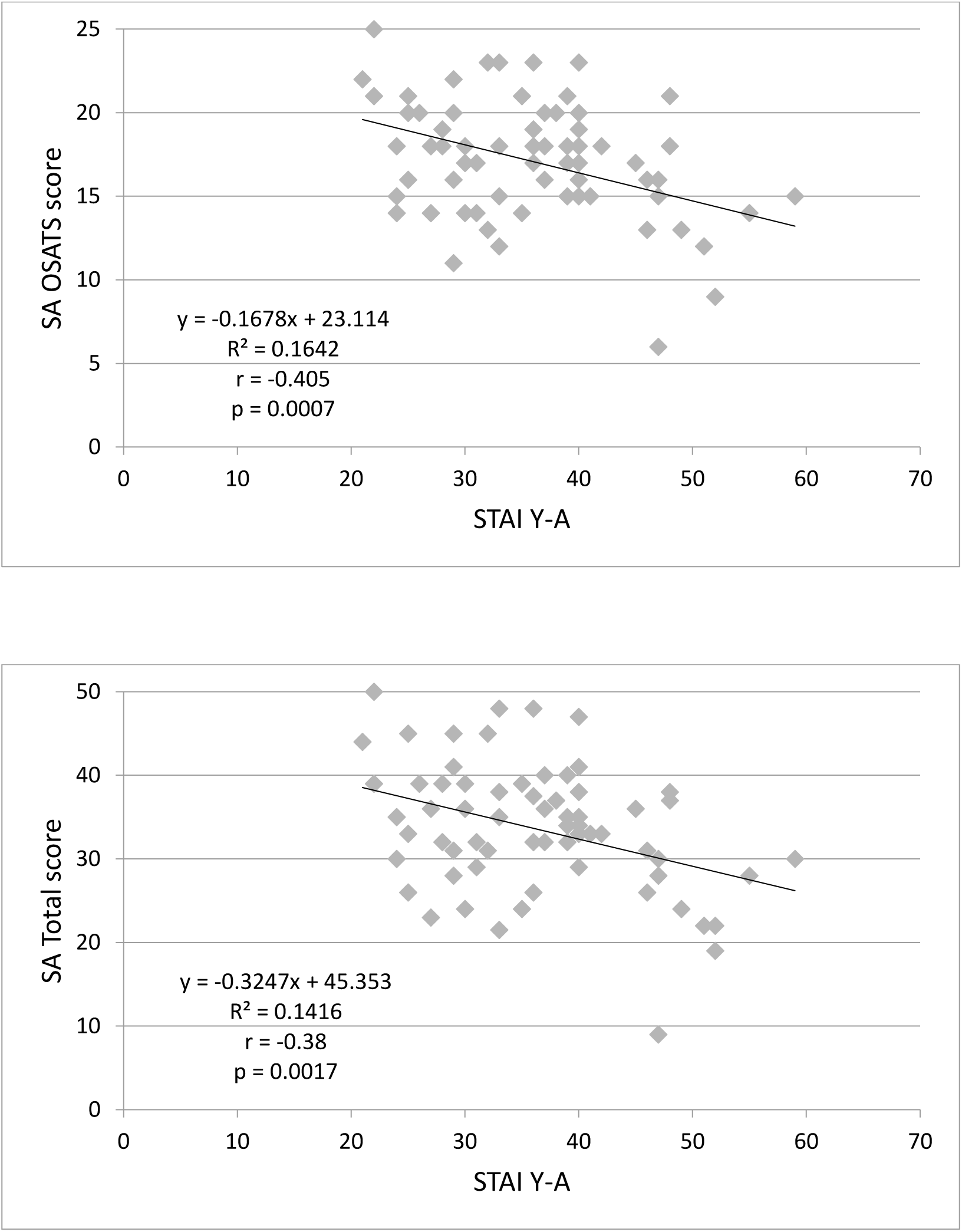
Analysis of correlation between the results of the STAI Y-A and the suture checklist, OSATS and total scores Legend: SA: self-assessment; EE: expert evaluation; r: correlation coefficient; R^2^: coefficient of determination; STAI-YA: State-Trait Anxiety Inventory

**Figure 3:**
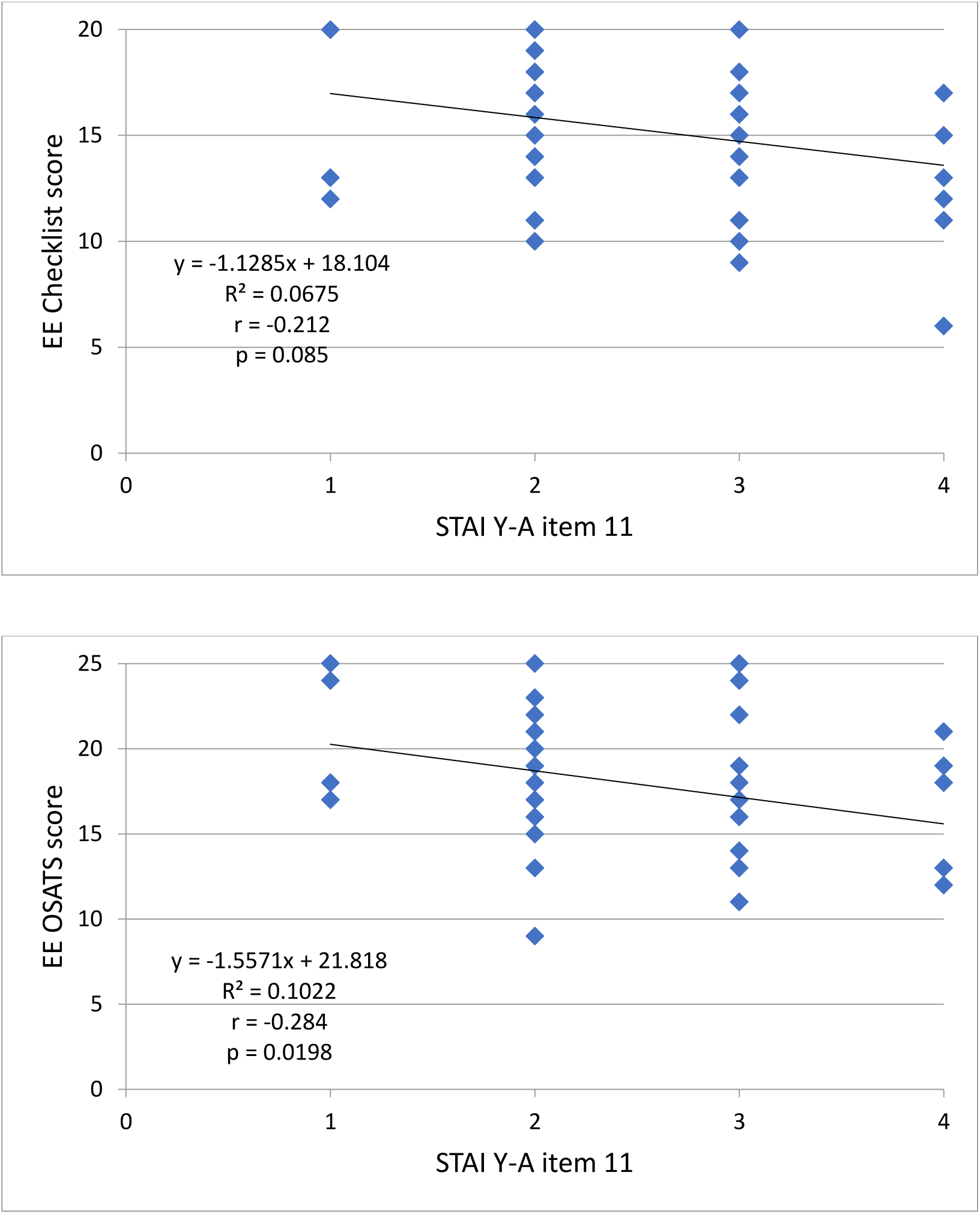
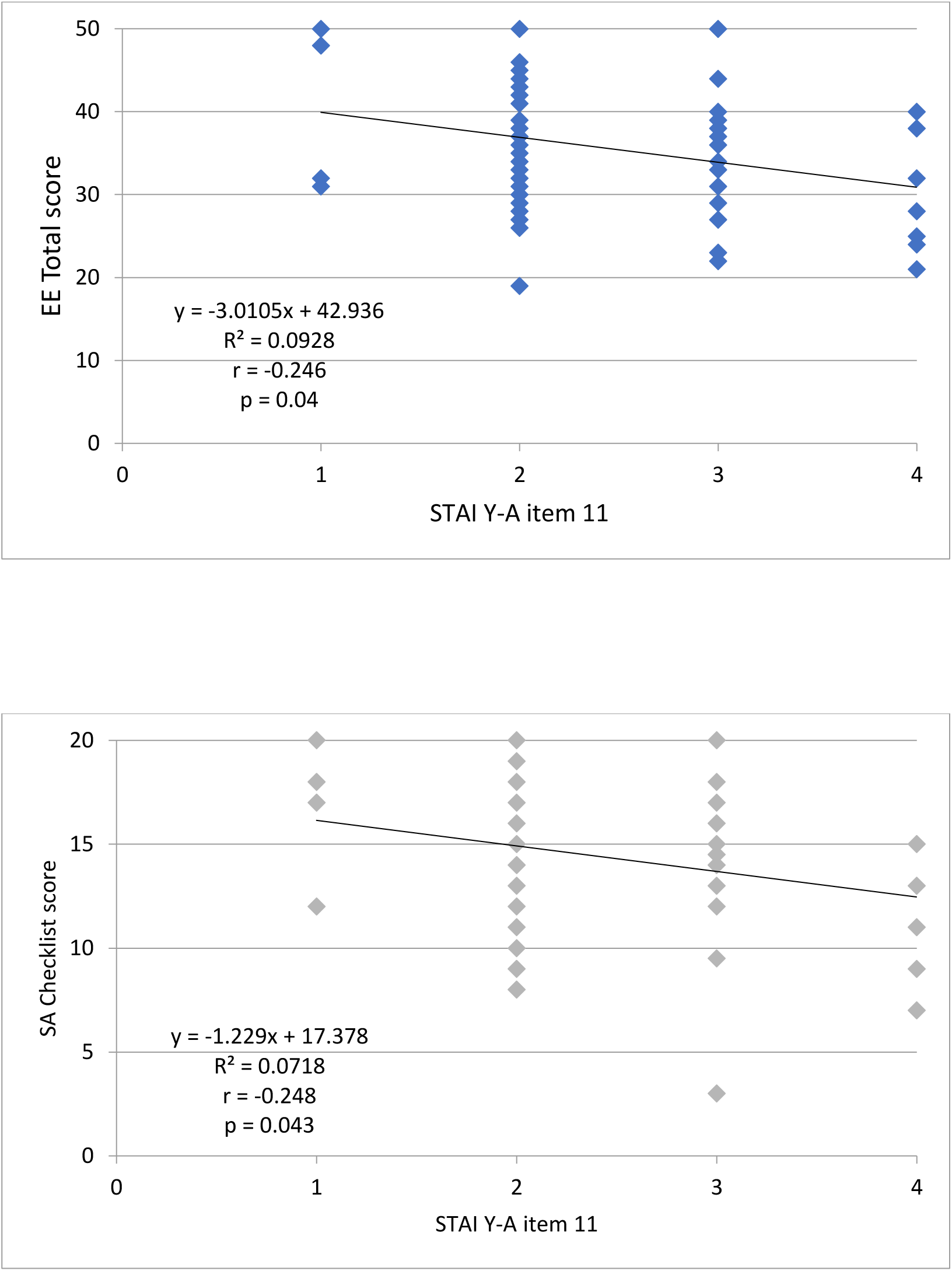
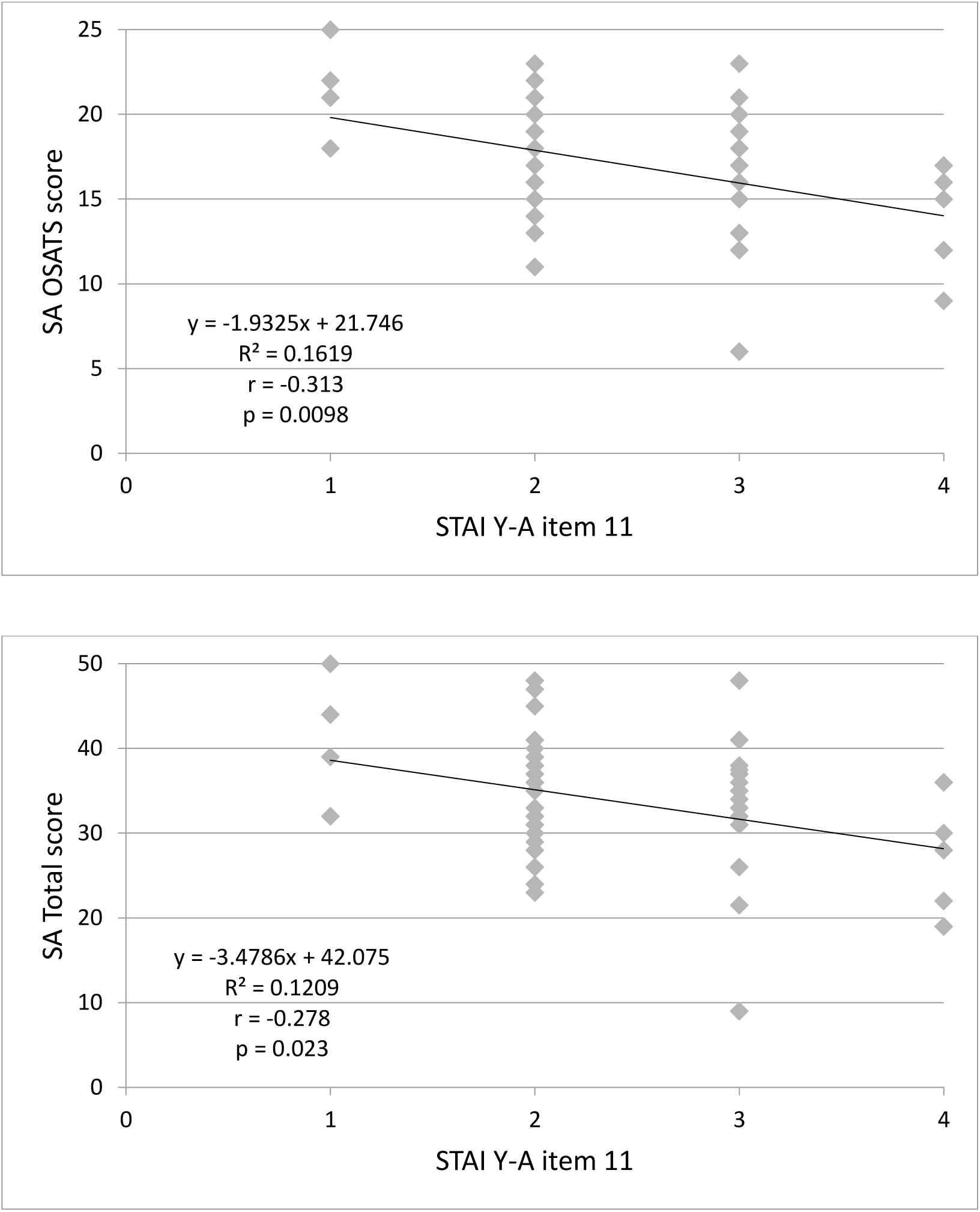
Analysis of correlation between the results of item 11 (self-confidence, ‘reversed’ item) of the STAI Y-A and the suture scores Legend: SA: Self-assessment; EE: Expert evaluation; r: correlation coefficient, R^2^: coefficient of determination. As a reminder, item 11 is reversed, and a score of 1 means high selfconfidence while a score of 4 means very low self-confidence.

The checklist score in EE was not correlated with the STAI Y-A score (r = -0.20; p = 0.096). However, there was a low but significant inverse correlation between the SA by checklist and the STAI Y-A scores (r = -0.29; p < 0.05). Similar results were found for the total score and the OSATS score, with a slightly stronger correlation between the level of anxiety and the SA OSATS score (r = -0.405; p < 0.001).

The checklist score was not correlated in EE with the self-confidence score (r = -0.21; p = 0.085), but it was correlated for the SA score (r = -0.248; p < 0.05). For all the other scores (OSATS and total), there was a significant inverse correlation (p < 0.05) between the EE, SA and self-confidence scores.

### Reasons for over- or underestimation

To refine the results, we studied extreme groups. The mechanisms underlying the over- or underestimation of skills were approached by analyzing the results for extreme groups (Figure 4), made up of, respectively, 28 trainee doctors in the ‘underestimation’ group and 12 trainee doctors in the ‘overestimation’ group. Their characteristics can be found in Table 2.

**Figure 4:**
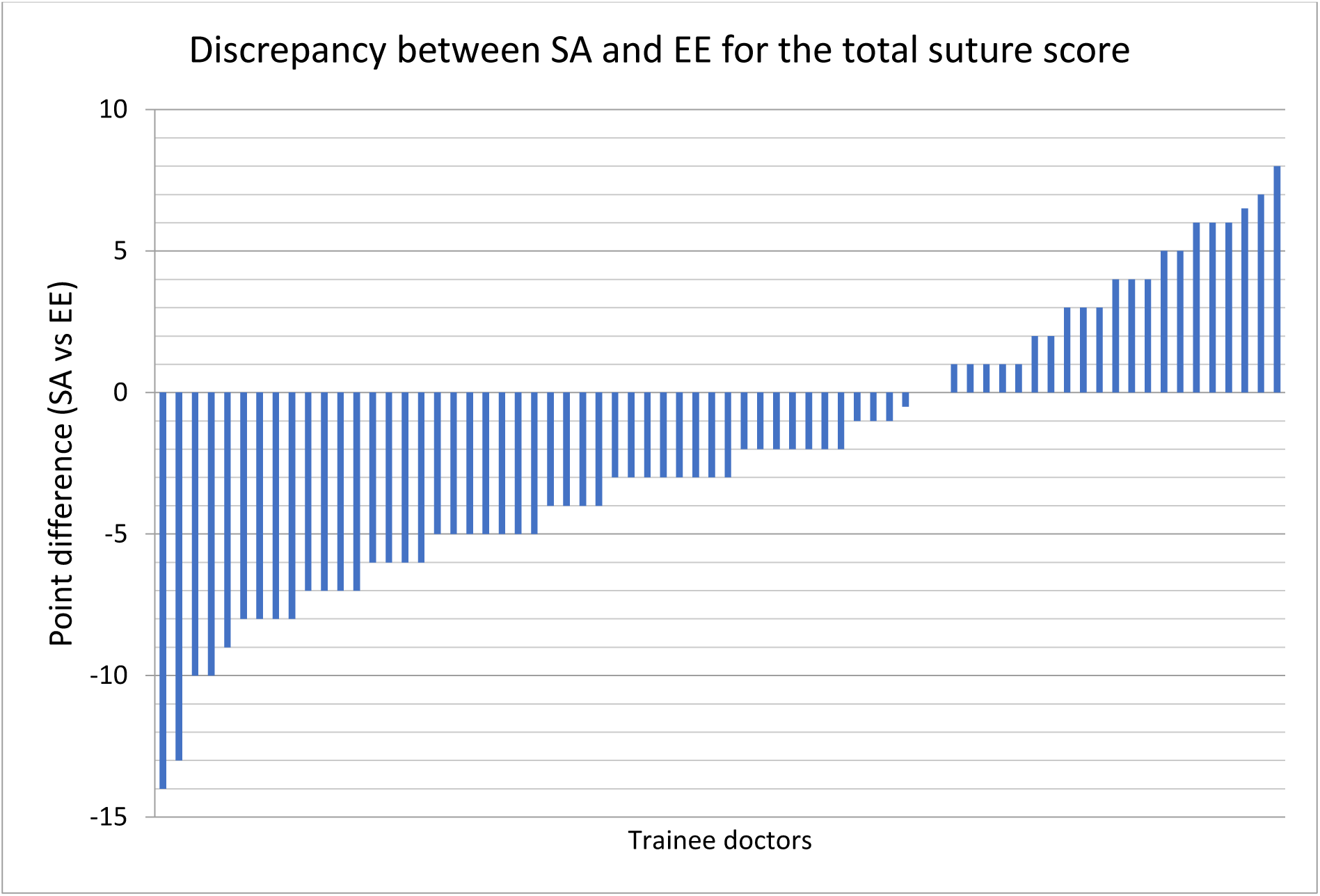
Analysis of ‘underestimation’ and ‘overestimation’ extreme groups Legend: SA: Self-assessment; EE: Expert evaluation; vs: versus

**Table 2:**
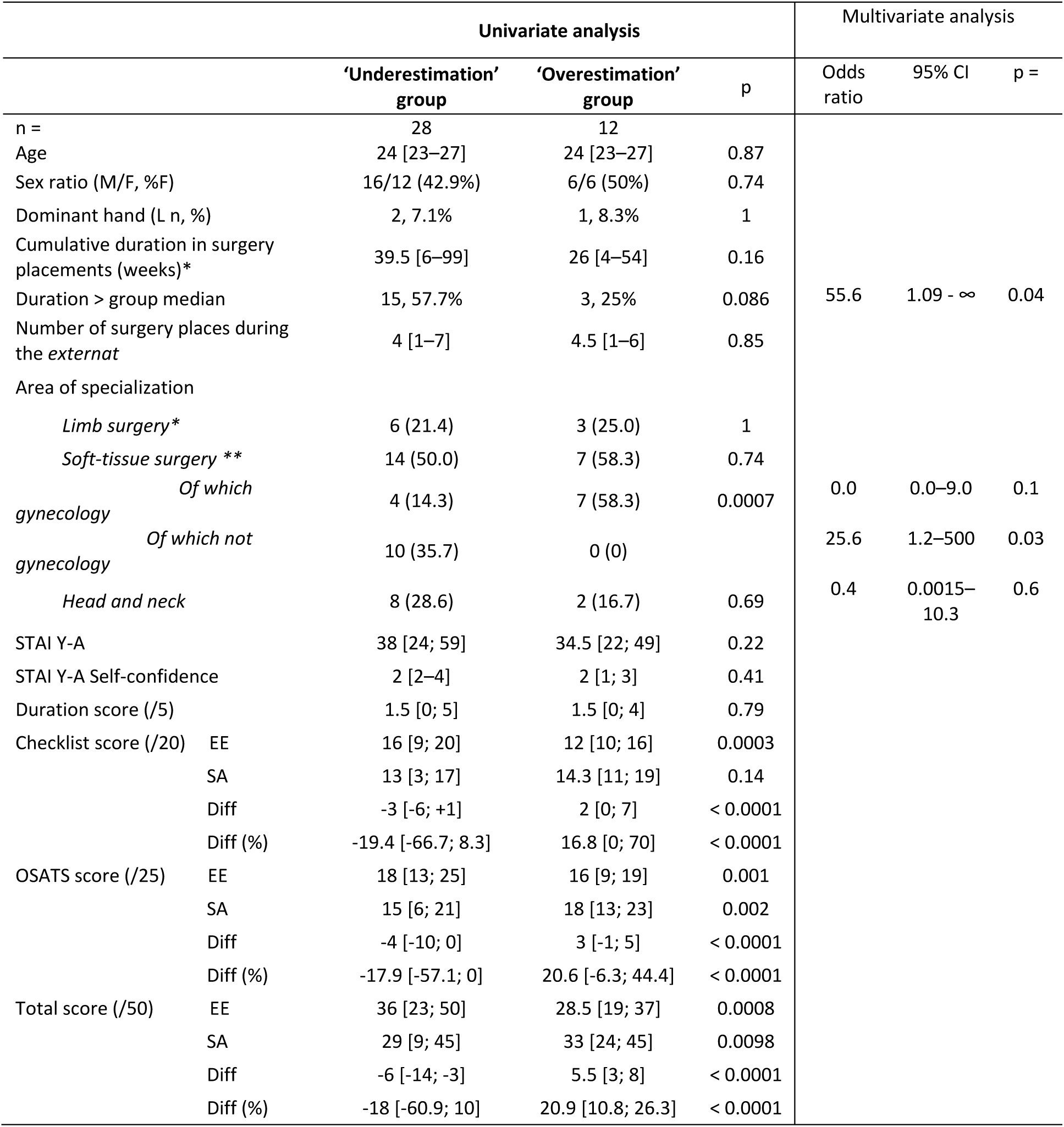
Characteristics of groups of trainees who over- or underestimate. Legend: SA: Self-assessment; Diff: difference in score between self-assessment (SA) and expert evaluation (EE), i.e.: Diff = SA score - EE score for the total suture scores, and the Checklist and OSATS sub-scores; Diff (%) = (SA score - EE score) / EE score*100; EE Expert evaluation; F: female; L: left; M: Male; min: minimum; max: maximum; Total score = OSATS score + checklist score + speed score; W = weeks; %: percentage N.B.: The cumulative duration of surgery placements reflects previous surgical experience. *orthopedics, vascular surgery; \*\**visceral surgery, urological surgery, pediatric surgery, gynecological surgery*; *** ENT, Maxillofacial surgery, Neurosurgery, eye surgery. *Quantitative variables are given as medians with extremes and qualitative variables are given as absolute values and percentages (%)*.

We did not find any differences between these two groups as regards demographic data or previous surgical experience. Trainee doctors specializing in gynecology tended to overestimate themselves, and significantly so (p = 0.005). The total, checklist and OSATS scores in EE given to the trainee doctors who overestimated themselves were lower compared to the trainee doctors who underestimated themselves, and significantly so (p < 0.001).

We performed a multivariate analysis to examine more thoroughly the determining factors relating to underestimation, including the area of specialization chosen, a long or short duration of surgical placements during the *externat*, and the STAI Y-A score (Table 2). Two factors were associated with over- or underestimating skills: the long duration of a placement and the learner’s specialization. A longer cumulative previous placement duration was inversely proportional to the risk of overestimation. This means that the less experience a learner had, the more they tended to overestimate their abilities. The second factor was that the nature of their specialization meant that trainee doctors specializing in soft-tissue surgery were 25 times more at risk of underestimating themselves than overestimating themselves.

## 4. Discussion

Our study on skin suturing identified several useful points that help to understand how self-assessment can be used as well as its limitations in the overall evaluation of learners who are also trainee surgeons.

Self-assessment seems to be a satisfactory evaluation technique as regards the OSATS and checklist scores. Combining three criteria in the evaluation seems to make self-assessments more reliable. However, and despite a very high correlation between the two assessments (SA and EE), it seems that learners have a general tendency to underestimate themselves.

In our study, we emphasized that the more anxious a learner was about the task at hand, the worse their SA results were (the OSATS score in particular) and the more they underestimated themselves. However, a learner’s anxiety had no effect on how they were assessed by an external examiner. This result shows clearly that, compared to the checklist score, the OSATS score is more subjective and therefore more linked both to one’s own representation of their expertise level and to psychological factors such as self-esteem and self-confidence, but also to the learner’s overall attitude, which includes their own self-confidence. On the other hand, the checklist grid is more objective and therefore less affected by the learner’s anxiety, which can definitely be useful for formative evaluation.

Anxiety could have severe negative consequences on cognition, decision-making, and learning. Studies conducted in the pediatric population have shown that learning new skills and demonstrating knowledge, through an assessment for example, can cause anxiety (Rappo *et al*., 2017; Raufelder & Kulakow, 2022; Westenberg *et al*., 2004). If a learner’s anxiety levels become too high, the learning process could be compromised because many classroom activities require using one’s working memory (Alloway, 2006) – which anxiety affects in a negative way (Moran, 2016). However, although anxiety affects performance in situations involving an evaluation, during exercises carried out at a later date the most anxious children seemed to have developed the same skills as their less anxious peers (Raghubar *et al*., 2010; Sotardi, 2018; Suárez-Pellicioni *et al*., 2016).

Stress and anxiety levels increase during medical studies (Lambert *et al*., 2020), especially for trainee surgeons who are exposed to complex care environments where tasks must be performed quickly and where various responsibilities, knowledge, and skills must be mastered in a short time. Yet stress and anxiety are associated with lower efficacy in healthcare and with higher patient morbidity and mortality (Arora et al., 2010). It is against this background that procedural simulation techniques were developed in healthcare. Procedure simulation allows for training in favorable conditions and in a safe environment, without putting patients at risk, and therefore without any unnecessary stress for the learner (Al-Elq, 2010; Antonoff *et al*., 2012; Cook, 2014; Liepert *et al*., 2019).

As regards the learner’s self-confidence and the validity of the SA taking this factor into account, in our study when learners had low self-confidence, they underestimated themselves, but examiners also had a tendency to give them lower grades. Lastly, apart from anxiety – which seems not to cause under-evaluation from the part of the assessor and therefore probably does not interfere with the learner’s procedural competences – high or low self-confidence, on the other hand, could interfere in the way in which an external assessor perceives a learner’s operational skills.

Self-confidence means feeling able to do something and believing that you have the ability and skills to fulfill an assignment or task. Self-confidence is about attitude, behavior, motivation, effort, and relationships with other people. An external assessor could, as a result of the learner’s self-confidence, become aware of other types of knowledge the learner has, especially soft skills such as behavioral competencies. In the case of learners with low self-confidence, their behavioral skills could be affected negatively through less accurate movements or hesitant behavior, for example, which could affect their overall ability to perform the procedure correctly. Although anxiety levels seem to increase during medical studies, so does self-confidence (Lambert *et al*., 2020). Simulation-based learning seems to be greatly effective in increasing self-confidence and reducing anxiety in trainees, which justifies the additional costs it entails compared to other teaching methods (Labrague *et al*., 2019; Yu *et al*., 2021).

Our findings also support the fact that the OSATS score could be more subjective and therefore more linked to the learner’s overall attitude, including their self-confidence. The OSATS score could also allow for a more comprehensive assessment of skills, covering expertise (knowledge), operational skills (know-how) and behavioral competencies (soft skills) (Abass *et al*., 2020; Reznick *et al*., 1997).

Three factors seem to affect whether a learner overestimates or underestimates themselves, namely (i) a lack of previous experience, creating a tendency to overestimate one’s abilities, (ii) the choice of certain areas of specialization, and (iii) a discrepancy in the SA and the EE. Trainees who overestimated themselves were found by the expert to be less competent than their colleagues.

Findings regarding previous experience confirm a theory put forward by David Dunning and Justin Kruger in 1999 (Kruger & Dunning, 1999), based on a 1871 hypothesis by Darwin: ‘Ignorance more frequently begets confidence than does knowledge’ (Darwin, 1888). The Dunning-Kruger effect (or ‘overconfidence effect’) is a cognitive bias in which people wrongly overestimate their knowledge or ability in a specific area. Novices are less aware of what’s expected from experts and might believe they have enough skills and knowledge from when they start learning.

Self-assessment is an innovative concept (McDowell, 1995; Siles-González & Solano-Ruiz, 2016) for learners and it forces them to reinterpret their own role. However, self-assessment also requires a sufficient amount of self-reflection and critical thinking, which falls under metacognition, defined as an awareness of one’s thought processes and the patterns behind them. Against this background, self-assessment helps learners to carry out their own evaluation of the knowledge and/or skills acquired during training. It also helps them to reflect on how their learning is progressing and to become aware of their strengths and weaknesses by encouraging them to work on the aspects they struggle with (Lafortune *et al*., 2000), which ultimately helps them to increase their chance of success by better showing the reasoning methods they use and by making it easier to acquire skills (Dubois, 2018), all the while increasing their self-confidence.

An assessment is a complex exercise that covers various approaches, procedures and goals based on different perspectives. To be valid and useful for learners, self-assessment must follow on from mentoring (Mcmahon, 1999; Siles-González & Solano-Ruiz, 2016). Generally speaking, taking part in the process of assessing one’s own skills helps to fully develop the ability to reflect on the knowledge acquired and to nurture critical thinking.

## 5. Conclusion

Lastly, it seems that a self-assessment grid can be used to evaluate learners. Checklist grids seem particularly well suited to formative self-assessment exercises as part of learning with a certain degree of independence because they seem to give less importance to subjective aspects than OSATS grids do. Yet while self-assessment grids seem applicable, vigilance is required in the case of some populations. The learner’s previous experience and choice of area of specialization should be discussed when interpreting the results of a self-assessment grid.

In addition, and in connection with the reduced duration of the initial medical course but also with the restricted duration of working time per week, many stakeholders argue that ‘bootcamps’ or seminars, such as the one organized by Angers University Hospital, could be promising alternatives to improve knowledge and skills but also to increase self-confidence and reduce stress levels in learners, especially at the start of junior doctor training (Esch *et al*., 2015; Lambert *et al*., 2020; Yeh *et al*., 2017). Formative self-reflection on learning, based on the use of self-assessment grids, could be combined with such programs to make them more effective.

## Data Availability

All data produced in the present study are available upon reasonable request to the authors or contained in the manuscript

## Notes

### Competing Interest Statement

The authors have declared no competing interest.

### Funding Statement

This study did not receive any funding

### Author Declarations

This study received approval by both Angers University Hospital Ethics Committee and CNIL (Commission Nationale Informatique et Libertes)(N 2019-002).

