## Supplemental File 1 for "Student self-assessment: feasibility, advantages and limitations Example of a workshop for trainee surgeons using a suture score"

Date:	Name of student:

Level of study:	Name of assessor:

**Description of the exercise:**

*On synthetic skin (Suture Skills Trainer 3500), perform a 4 cm linear intradermal continuous suture, starting with an intradermal reverse cutting suture by hand and ending with a knot tied with a needle holder (intradermal or outside the wound, at the learner’s discretion).*

**Objectives:**

- Speed score: Operative time under 5 minutes.
- Suture and knot scores > 15/20
- Overall scale > 20/25

**(1) Speed score: _____/ 5**

*(subtract 1 point per minute beyond 5 minutes, i.e. 4 points if completed between 5’01 and 6’00, 3 points if completed between 6’01 and 7’00, etc. until no points are left. No negative numbers)*

Duration of the exercise _______ minutes and _________ seconds

**(2) Checklist score:** (one point per correctly performed item) **_________ / 20**

***Score for completing the starting knot by hand: ___ / 7***

□ Places the needle in the needle holder correctly (~ perpendicular to the holder and at ~ 2/3 of the distance from the needle tip)

□ Places the suture deep in the wound to bury the knot

□ Both ends of the suture thread should arrive on the same side of the thread loop

□ Places the tip in the dermis correctly

□ Tightens the knot without excessive tension

□ Makes at least 4 loops and knows how to make knots in both directions

□ Cuts the blind end of the thread at a short distance from the knot

***Suture score:____/ 7***

□ Completes the continuous suture in the “correct direction” (from top to bottom, or laterally starting at the side of the dominant hand)

□ Handles the wound edges gently and does not cut through the skin

□ Inserts the needle parallel to the skin surface, following the curve of the needle

□ Passes the needle through the tissue using supination and removes the needle using pronation

□ Places bites in equal depth and so that they emerge out of the skin from the previous exit point, without crossing the thread

□ Bites are equal in size and regular

□ The skin edges should be in apposition, with neither too much or too little tension, and with no scar with a ‘rail track’ appearance

***Score for laying a knot using a needle-holder: __/ 6***

□ Knows how to use the continuous suture thread to create a loop and lay an end knot

□ Starts the knot with a double loop

□ Subsequent loops are inverted compared to the first ones

□ Makes at least 4 loops, alternating directions

□ Tightens the knot without excessive tension on the continuous suture

□ Cuts the thread without excess length

**(3) Overall assessment scale:**

For each line, circle the number that best corresponds to the performance assessed, without taking the participant’s level into account.

*In this assessment, the learner starts with 13/25 points. For each item below, add or subtract points as appropriate.*

| **Knowledge and handling of instruments** | | | | | |
| --- | --- | --- | --- | --- | --- |
| -2  Cannot name the instruments, chooses the incorrect instrument, handles instruments in an inappropriate manner | | -1 | 0  Can name only some instruments, hesitates or changes his or her mind before picking an instrument up, handles instruments correctly most of the time | +1 | +2  Can name all the instruments correctly, chooses the appropriate instrument easily, uses instruments correctly in all circumstances |
| **Quality of the suture** | | | | | |
| -2  Poor technique, poor dexterity, very irregular skin suture | -1 | | 0  Average technique, good dexterity, suture looks acceptable | +1 | +2  Excellent technique, excellent dexterity, perfect suture |
| **Quality of knots** | | | | | |
| -2  Poor technique, does not make at least 4 loops, incorrect knots (too tight, blocked, too loose, at risk of becoming undone, etc.) | -1 | | 0  Average techniques, some knots tied better than others, most knots tightened correctly | +1 | +2  Excellent technique, knots tied very well, knows completely secure |
| **Tissue handling** | | | | | |
| -2  Often uses too much force in his or her movements, damages the tissue | -1 | | 0  Handles tissue carefully but damages it slightly | +1 | +2  Handles tissue very carefully, with no or minimal damage to the tissue |
| **Movements** | | | | | |
| -2  Many unnecessary movements, often stops and starts again, often handles the tissue with the instruments more than once | -1 | | 0  A few unnecessary movements but reasonably efficient, smooth progression, occasionally handles the tissue more than once | +1 | +2  Keeps movements to a minimum, very good flow and steady pace, minimal gripping of the tissue |
| **Overall performance** |  | |  |  |  |
| -2  Failed to perform procedure, skill not acquired | -1 | | 0  Procedure performed correctly, skill acquired | +1 | +2  Procedure performed perfectly, above-average skill |

**Total score** (sum of each item)**: ________ / 25**

**Multimodal assessment grid - Total score ( 1 + 2 + 3) = _____ /50**
